# Multisensory Integration of Naturalistic Speech and Gestures in Autistic Adults

**DOI:** 10.1101/2024.06.22.24309300

**Authors:** Magdalena Matyjek, Sotaro Kita, Mireia Torralba Cuello, Salvador Soto Faraco

**Affiliations:** Center for Brain and Cognition, Universitat Pompeu Fabra, Spain; Humboldt-Universität zu Berlin, Germany; University College London, UK; University of Warwick, UK; Institució Catalana de Recerca i Estudis Avançats (ICREA), Barcelona, Spain

**Author notes:** **Competing Interest Statement** The authors declare no competing (financial or otherwise) interests. **Funding statement** This project has received funding from the European Union’s Horizon 2020 research and innovation programme under the Marie Skłodowska-Curie grant agreement no 945380, DAAD PRIME Fellowship, Agencia Estatal de Investigación grant PID2019-108531GB-I00 AEI/FEDER, and AGAUR Generalitat de Catalunya grant 2021 SGR 00911.

**Keywords:** Autism, multisensory integration, EEG, iconic gestures, audio-visual speech

## Abstract

Seeing the speaker facilitates auditory speech comprehension through audio-visual integration. This is especially the case in challenging listening conditions, such as in real-life social environments. Autism has been associated with atypicalities in integrating audio-visual information, potentially underlying social difficulties in this population. The present study investigated multisensory integration (MSI) in speech processing among autistic (N=35) and neurotypical (N=35) adults. Participants performed a speech-in-noise task in a realistic multispeaker social scenario with audio-visual, auditory, or visual trials while their brain activity was recorded using EEG. The neurotypical group demonstrated a non-linear audio-visual interaction in alpha suppression, whereas the autistic group showed merely additive processing of these same inputs. Despite these differences in neural correlates, both groups achieved similar behavioural audio-visual facilitation outcomes. These findings suggest that although autistic and neurotypical brains might process multisensory cues differently, they achieve comparable benefits from audio-visual speech. These results contribute to the growing body of literature on MSI atypicalities in autism.

## 1. Introduction

Face to face conversations, like many social interactions, rely on the processing and integration of auditory information (such as acoustic, phonetic, and phonological cues), as well as visual information (such as lip movements, facial expressions, and body gestures. These cues collectively inform various aspects of communication, including lexical access, syntax, semantics, and pragmatics (Brunellière et al., 2013; Brunellière & Soto-faraco, 2015). In the neurotypical^1^ populations, the integration of congruent visual and auditory information increases communication efficiency and speech perception, particularly in adverse listening conditions (Bernstein et al., 2004; Ross et al., 2007; Schepers et al., 2013; Sumby & Pollack, 1954). Given the critical role of multisensory integration (MSI) processes in development, it is not surprising that alterations in MSI have been associated with certain neurodevelopmental conditions or variations, such as autism spectrum disorder (hereafter, autism) (Wallace et al., 2020).

Autism, characterised by difficulties in social interactions/communication, repetitive behaviours, and sensory atypicalities (American Psychiatric Association, 2013), shows differences from the neurotypical development in the processing of single modality information and in the integration of multisensory signals (Iarocci & McDonald, 2006; Koelewijn et al., 2010; Marco et al., 2011; Stevenson, Segers, et al., 2014; Zhang et al., 2019). Furthermore, evidence suggests that these atypicalities may be particularly pronounced in the context of social cues, like speech (Stevenson, Siemann, et al., 2014). While existing literature has provided valuable insights into MSI in autism, existing research has often used well-controlled and idealised stimuli, far from the complexity of typical social contexts. Therefore, there remains a gap in understanding how MSI processes play out in more naturalistic social settings. Here, we aim to address this gap by investigating behavioural and neural MSI in autistic^2^ and neurotypical adults using audio-visual stimuli embedded in a naturalistic, linguistic, pragmatic, and social context to gauge MSI in more ecologically valid situations.

In particular, social interactions are not only dynamic and perceptually complex, but also multisensory in nature and unfold in parallel with other cognitive processes, like attention (Soto-Faraco et al., 2019). To consider these elements in our design and improve real-world generalisation, we prioritised ecological validity (Maguire, 2012; Matusz et al., 2019). In particular, we used an environment with a rich socio-linguistic context: three persons playing a word game in an online video-chat facilitated by a teacher (this paradigm has previously been validated by our group; (Matyjek et al., 2024)). This setting provides natural context-based pragmatics and semantic priors for meaningful speech stimuli, which are complete words (as opposed to isolated syllables or out-of-context utterances; (Stevenson, Siemann, et al., 2014; Williams et al., 2004)). Further, we also included a full view of the actors’ torso and face, because MSI benefit relies on multiple naturally produced *visual articulators* (Drijvers & Özyürek, 2017), including orofacial movements, head and eyebrows dynamics (Thomas & Jordan, 2004; Yehia et al., 2002). Finally, in real-life conversations, hand gestures are well coordinated with speech (Goldin-Meadow & Wagner, 2005; Kita & Özyürek, 2003; McNeill, 1992; Özyürek et al., 2007). *Iconic* gestures (typically hand-produced movements representing characteristic actions or attributes; (Kita & Özyürek, 2003)) are endowed with semantic information (Drijvers & Özyürek, 2017). Prior research addressing integration of gestures with speech in neurotypical populations show improvements in speech comprehension and characteristic neural effects (Drijvers & Özyürek, 2017; Holler et al., 2014; Özyürek, 2014). As is the case with audio-visual MSI, this gestural benefit is especially visible in adverse listening conditions (Drijvers, Vaitonytė, et al., 2019; Drijvers, van der Plas, et al., 2019; Drijvers & Özyürek, 2017; Holle et al., 2010; Özyürek, 2014).

Previous studies of audio-visual and speech-gesture integration in autism have focused mainly on children and adolescents, and their results are mixed. Among the audio-visual speech integration literature, some studies suggest atypicalities in single-modality processing (Williams et al., 2004), others point to specific differences in MSI processing (Silverman et al., 2010; Stevenson, Siemann, et al., 2014), some report atypicalities in both unimodal and MSI processing (Foxe, Molholm, Bene, et al., 2015; Irwin et al., 2011, 2011; Russo et al., 2010; Smith & Bennetto, 2007; Stefanou et al., 2020; Stevenson et al., 2017), and some, based on behavioural outcomes, cannot clearly identify which process is responsible for group differences (Alcántara et al., 2004; Bebko et al., 2006). Despite possible atypicalities in early life, there is evidence for developmental improvement of MSI in autism (Beker et al., 2018; Brandwein et al., 2013; Feldman et al., 2018; Foxe, Molholm, Bene, et al., 2015; Foxe, Molholm, Del Bene, et al., 2015), which may lead to typical integration performance – at least at a behavioural level – in adulthood (Keane et al., 2010; van der Smagt et al., 2007). The speech-gesture integration findings in autism also show inconsistencies. Silverman et al. (2010) observed that while co-speech gestures speeded up gaze to targets vs. distractors in neurotypical adolescents, the same gestures *delayed* focusing the gaze on the targets in their autistic peers. Nevertheless, the gaze outcomes were an implicit measure and there were no statistically significant differences between the groups in the task that participants were instructed to perform (reaction times of clicking on the drawing corresponding to the description heard in the audio). Thus, the gestural effect in relation to autism is not clear in this study. More recent studies reported that autistic children (Dargue et al., 2021; Kurt, 2011) and adults (Mazzini et al., 2023) benefit from co-speech gestures in narrative recall, acquiring receptive language skills, and speech comprehension. For example, (Mazzini et al., 2023) showed that the behavioural benefit from gestures in degraded speech is of comparable size in both autistics and neurotypicals. Together, given the mounting evidence for general MSI atypicalities in autism, semantically laden gestures (like iconic gestures) seem to have the potential to facilitate otherwise atypical processing of semantic multisensory information in autism.

From the literature above, it is plausible that variations in the neural markers of MSI in autism are not necessarily accompanied by behavioural alterations, especially towards adulthood. For example, (Stefanou et al., 2020) found no group differences in behaviour between autistic and non-autistic adolescents when they were asked to quickly respond to auditory, visual, or audio-visual non-social targets. However, they observed significant neural differences between the groups, including delayed and spatially constrained onset of MSI neural markers in autism. This dissociation has been shown also in neurotypicals: neural MSI effects have been reported in the absence of the behavioural multisensory gain (Santangelo et al., 2008). Hence, integrating both behavioural and neural assessments is essential for a more comprehensive characterisation of MSI in general, and especially in autism. Oscillatory dynamics in the EEG signal can index the integration of complex information across modalities (Varela et al., 2001), including MSI (Senkowski et al., 2008), and may be less susceptible to variability when using complex stimuli (Matyjek et al., 2024). This is especially important when using more ecologically valid stimuli, which often sacrifice experimental control and signal to noise ratio. For instance, alpha/beta suppressions and gamma power increases have been functionally linked to the enhancement of speech comprehension through gestures (Drijvers et al., 2018b, 2018a).

In the present study, we asked how multisensory speech is processed and integrated in autism under complex, naturalistic social conditions. To this end, we measured behavioural (accuracy of word recognition) and neural (alpha suppression) responses of autistic and non-autistic adults, while they were watching a video conference with multiple actors playing a word game. The actors took turns naming actions which could be performed in a given situation (e.g., in a restaurant), using corresponding iconic gestures. In unimodal trials either the actor’s camera was turned off (auditory only) or the audio was too noisy to understand speech (visual only). In bimodal trials, the actors could be both seen and heard. All trials included pink noise to ensure sufficiently adverse listening conditions.

We expected that while the behavioural benefit from speech-gesture MSI is similar in autistic (AUT) and neurotypical (NT) adults (Mazzini et al., 2023), multisensory integration is realised differently at the neural level (linked to less suppressed alpha) in autism. Specifically, we predicted that:

1) AUT would show a bimodal benefit at a behavioural level, as measured with speech perception accuracy (better accuracy in AV than in A and V).
2) The NT and AUT groups would show similar benefit in bimodal in comparison to unimodal trials in terms of behaviour (here: similar AV-max(A,V) difference in accuracy in AUT and in NT).
3) AUT would show the MSI effect of bimodal vs unimodal speech at the neuronal level, as measured with alpha suppression (stronger alpha suppression in the AV trials than in the sum of A and V trials).
4) AUT would show a smaller MSI effect than NT at the neuronal level (less alpha power suppression).

The hypotheses and analyses are preregistered at https://osf.io/4765g.

## 2. Materials and methods

### 2.1. Sample size calculation and power analysis

To determine the sample size for this study, we based the effect size on a meta-analysis of audio-visual multisensory integration in autism yielding Hedge’s g of 0.41 (Feldman et al., 2018). We performed a power analysis with the *pwr* package in R (Champely, 2020). A multiple regression power calculation with effect size f^2^=0.205 (equivalent of g=0.41), 80% power, alpha level of 0.05, and 2 numerator degrees of freedom (2 groups – 1, times 3 conditions −1) yielded a sample size of N=25. However, we anticipated that our true effect size may be smaller given the naturalistic nature of our stimuli. Therefore, we planned to collect data from 35 participants per group, which would allow us to detect a medium-large effect size of f^2^=0.14.

### 2.2. Participants

The characteristics of the autistic and non-autistic participants are summarised in Table 1. All participants gave written consent to participate in the study. The study protocol and data handling were approved by the Institutional Committee for Ethical Review of Projects at University Pompeu Fabra (CIREP-UPF). All the participants were Spanish-speaking volunteers between the age of 18 and 60, with no intellectual disability, and with normal or corrected-to-normal vision and audition.

**Table 1.**
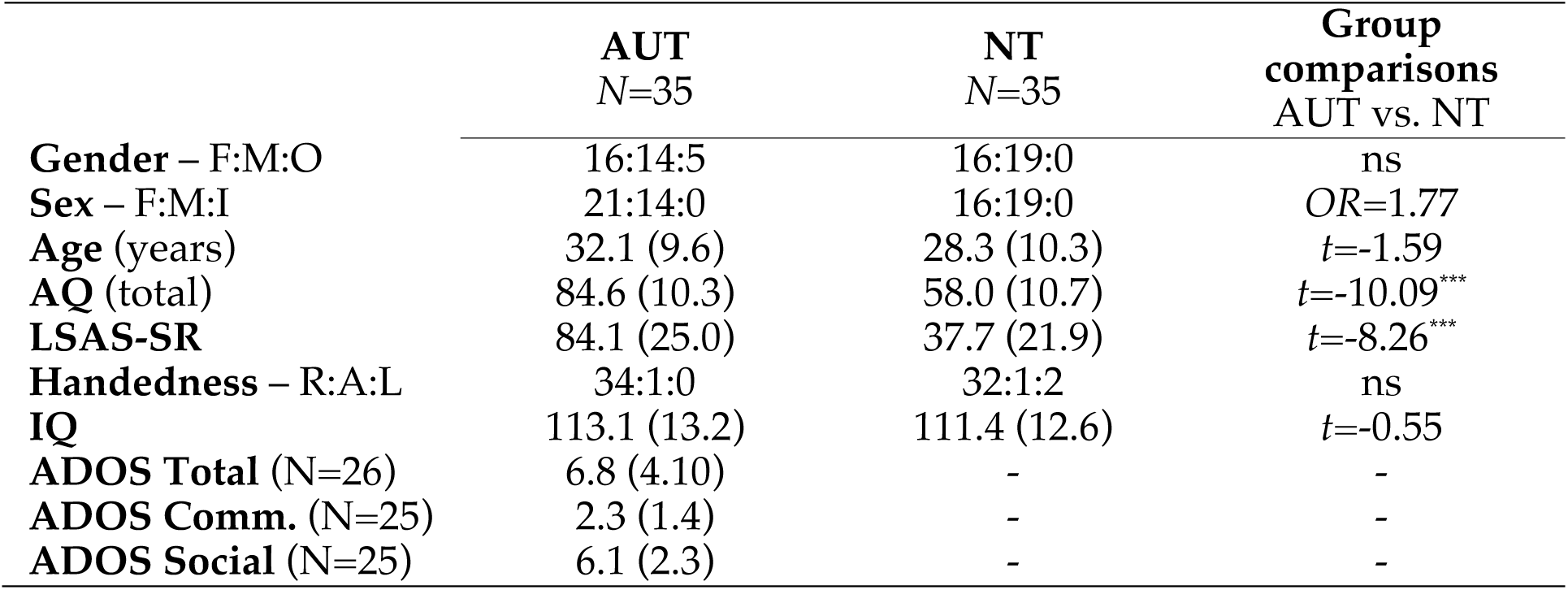
Demographic and trait characteristics of subject samples in all groups. Count is provided for gender, sex, and handedness, and means (with standard deviations) for all other items. F/M/O/I=female, male, other, intersexual, AQ=Autism Spectrum Quotient 10-item Spanish version (Lugo-Marín et al., 2019), LSAS-SR=Liebowitz Social Anxiety Scale – Self Reported, R/A/L=Right, Ambidextrous, Left, IQ=Intelligence Quotient measured with Raven’s Progressive Matrices 2 (John & Raven, 2003), OR=odds ratio in Fisher’s Exact Test. Statistically significant tests were marked with *** for p<.001. *Ns*=non-significant.

|  | **AUT**<br>N=35 | **NT**<br>N=35 | **Group**<br>**comparisons**<br>AUT vs. NT |
| --- | --- | --- | --- |
| **Gender** – F:M:O | 16:14:5 | 16:19:0 | ns |
| **Sex** – F:M:I | 21:14:0 | 16:19:0 | OR=1.77 |
| **Age** (years) | 32.1 (9.6) | 28.3 (10.3) | $t=-1.59$ |
| **AQ** (total) | 84.6 (10.3) | 58.0 (10.7) | $t=-10.09^{***}$ |
| **LSAS-SR** | 84.1 (25.0) | 37.7 (21.9) | $t=-8.26^{***}$ |
| **Handedness** – R:A:L | 34:1:0 | 32:1:2 | ns |
| **IQ** | 113.1 (13.2) | 111.4 (12.6) | $t=-0.55$ |
| **ADOS Total** (N=26) | 6.8 (4.10) | - | - |
| **ADOS Comm.** (N=25) | 2.3 (1.4) | - | - |
| **ADOS Social** (N=25) | 6.1 (2.3) | - | - |

Autistic (AUT) participants were recruited through invitations shared by collaborating autism centres in Barcelona, social media, flyers at public events, and word of mouth within the local autism community. All AUT participants had a confirmed diagnosis of autism spectrum disorder made in adulthood by professionals in specialised autism centres, in accordance with DSM-5 criteria (American Psychiatric Association, 2013). Of these, diagnosis was confirmed by the Autism Diagnostic Observation Schedule (ADOS; Lord et al., 2000) in 26 participants, and by the Autism Diagnostic Interview-Revised (ADI-R; Bölte & Poustka, 2001; Lord et al., 1994) in 14 participants. Co-occurring diagnoses in the AUT group: attention deficit (hyperactivity) disorder (N=7), depression (N=16), and anxiety (N=17). Additional diagnostic information can be found in Supplementary Material (section 1).

Data from 39 neurotypical (NT) participants were collected in an earlier study with identical design and method (Matyjek et al., 2024) and were re-used here as a comparison group. Note that while these data had already been analysed, none of the hypotheses in the current study are dependent on the effects observed there. To match the autistic (AUT) and neurotypical (NT) groups on count, biological sex, age, and IQ, we removed four NT datasets of the most extreme age and IQ scores (before analysing the behavioural or EEG data), resulting in the final group of 35 NT participants. None of the NT participants had a history of neuro-psychological or neuro-psychiatric disorders. All were recruited via the Center for Brain and Cognition (University Pompeu Fabra) participant database.

### 2.3. Task and stimuli

The task used was identical to that described in detail in Matyjek et al. (2024). An overview of a trial is shown in Figure 1. In short, participants watched a (simulated) recording of a videoconference with three actors playing a word game (see a demo video of the paradigm at https://osf.io/kdj48). The actors uttered verbs matching a given situation (e.g., “What do we do in a restaurant?” -“We drink.”) using co-occurring iconic gestures (e.g., a hand approaching the mouth as if drinking from a cup). Three conditions were introduced: bimodal audio-visual (AV; actors can be both heard and seen), unisensory visual (V; no audio speech signals, as if the actor’s microphone was malfunctioning) or auditory (A; no visual signals as if the camera was “turned off”). Pink noise was added to each trial to create adverse listening conditions (audio speech to noise ratio=0.44). 61 verbs were used, each presented in the three conditions (183 trials). The stimulus videos were preceded (random presentation time between 0.3-0.7 s) and followed (0.2-0.3 s) by a fixation symbol (“video loading” circle). The task was to indicate the target verb uttered by the actor by pressing one of four keys on a computer keyboard corresponding to: the correct word, a semantically similar word, a phonetically similar word, and an unrelated word (locations were randomised). The words were presented on the screen until a response was given or until the maximum time of 3 s elapsed.

**Figure 1.**
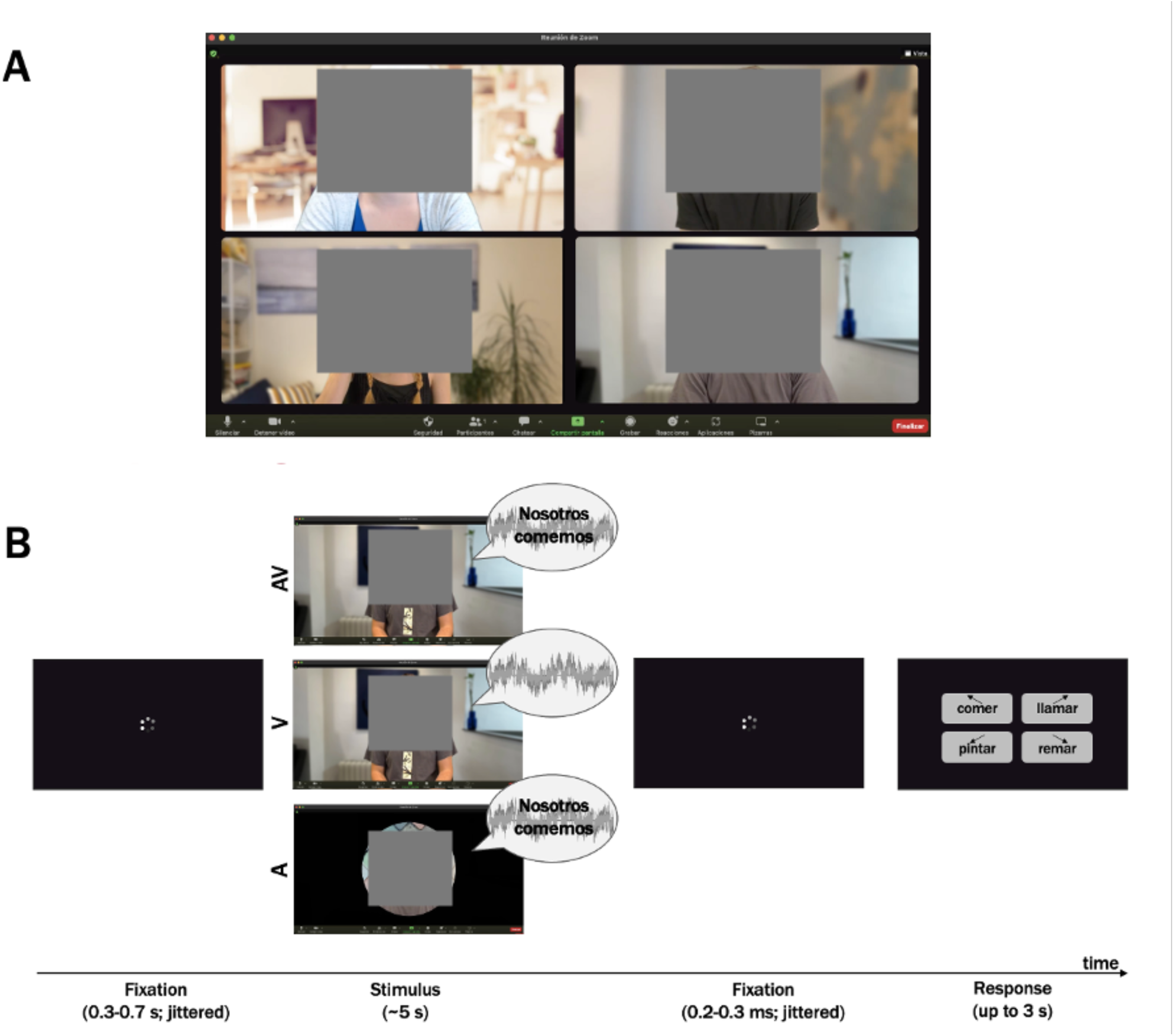
(A) Social situation: a videoconference with the teacher (top left) moderating a word game for three participants. (B) Schematic representation of the trial order. The grey waves in the speech bubble indicate pink noise to create an adverse listening condition. Grey squares have been added for anonymity according to MedRxiv policy, but the stimuli used in the experiment featured nnoccluded view of the speaker.

Each target verb appeared once per condition and per actor, and the same modality condition (AV, A, V) was not presented more than three trials in a row. The order of the words was pseudo-randomised (by using three versions of the trial order with orthogonal assignments of condition for each word, and a different order of words balanced across participants). The task started with a training which was repeated until the instructions were clear to each participant. To promote divided attention to both sensory channels in the task, participants were asked to monitor for occasional (N=19) glitches (“catch trials”) in either the visual or the auditory channel (frozen frames or a burst of noise in the microphone, respectively). The detailed description of the stimuli, target verbs, and the elements of the design ensuring an embedded social context can be found in Matyjek et al. (2024).

### 2.4. EEG data acquisition and processing

The continuous EEG signal was acquired using the Brain Vision Recorder at a sampling rate of 500 Hz, with data collected from 61 active electrodes (actiCHAmp; Brain Products, Gmbh). Electrodes were positioned based on the extended 10–10 international electrode placement system and secured onto an elastic cap, with AFz serving as the ground. Impedances were maintained below 20 kΩ. To record eye movements, additional electrodes were positioned at the outer canthi of the right eye (HEOG) and below the left eye (VEOG). During recording, the online reference was set at the tip of the participant’s nose. Offline pre-processing of the EEG signal was conducted using Brain Vision Analyzer software (Brain Products). First, we applied a 0.1 Hz high-pass Butterworth filter (zero phase shift order 8) to all data (no low-pass filter was applied). Visual inspection was used to detect channels with poor signal quality (those that showed a flat signal (detached electrode) or that presented fluctuations in the voltage (standard deviation) larger than the rest of electrodes of the set) were interpolated based on the neighbouring channels using spherical splines of order 4 (2.08% of all channels). Subsequently, average reference was performed. Continuous data were divided into segments ranging from −1000 ms before to 3000 ms after the acoustic onset of the target verb. Local DC detrending was applied to segments to remove slow drifts in the baseline, and those containing large muscular artifacts were removed using a semi-automatic procedure. An independent component analysis algorithm (restricted fast ICA) was used to identify and correct activity related to blinks and eye movements from the data, resulting in an average removal of 2.81 components per subject. Manual artifact rejection was then performed on the remaining segments. The number of artifact-free segments did not significantly differ across experimental conditions, *F*(2,13.40)=0.81, *p*=.45, and group*condition interaction was not significant, *F*(2,6.30)=0.38, *p*=.68, but there was on average one segment more in NT than in AUT, *F*(1,69.72)=8.42, *p*=.004 (the mean number of segments left for groups and conditions were: AV_AUT_=58.17, A_AUT_=58.26, V_AUT_=58.37, AV_NT_=58.86, A_NT_=59.51, V_NT_=59.89). Finally, segments were divided into AV, A, and V trials for subject-level and grand averaging. The processed data were exported to Matlab for further processing using the FieldTrip toolbox (Oostenveld et al., 2011).

### 2.5. Data analysis

All statistical analyses were conducted in R v. 4.3.2. (R Core Team, 2023). Alpha level was set to .05. For all models, treatment contrasts were used with AV set as the reference level. Note that predictions 1 and 3 pertain to AUT data only, as the data including all NT participants in this study were already analysed and reported elsewhere (Matyjek et al. 2024). Nevertheless, we also report the NT data here for completeness.

#### 2.5.1. Accuracy

For the analysis of word recognition accuracy in AUT (prediction 1), responses were considered correct when the participant chose the word option matching the word uttered by the actor in that trial. Choosing one of the three other words or failing to respond within the time limit (3 s) resulted in an incorrect response. A generalised linear mixed model (GLMM) was built with a binary outcome variable with single-trial correct (=1) and incorrect (=0) responses, condition (AV, A, V) as a predictor, and random intercepts and slopes for conditions within each subject’s ID and for conditions within each verb.

For the analysis of behavioural MSI benefit (to test prediction 3), we used the equation: *AV* − max(*A*, *V*) for each participant, where AV is the percent of correct responses for this participant in AV trials, and max(A,V) is the highest percent of correct responses for this participant between A and V trials. Then, these behavioural MSI benefit scores were compared between the groups with a two-sided t-test.

#### 2.5.2. Reaction times

Given that we have recorded reaction times (RTs), we entered them in an exploratory analysis. Please note that RT may be less sensitive to MSI effects in our design, because of the delayed response prompt introduced to avoid motor contamination in the EEG signal. For this reason, we pre-registered an exploratory approach for the RT analysis. RTs faster than 200 ms and outliers (RTs outside the range median +/-2 x median absolute deviation; Leys et al., 2013) were removed. Additionally, RTs corresponding to trials in which the EEG data were rejected due to the presence of artifacts were also removed. We built a GLMM with condition, group, and their interaction as predictors. For pairwise comparisons within the model, Holm correction was administered via the *glht* function from the *multcom* R package (Hothorn et al., 2008). To estimate the main effects of the multilevel predictor condition, a type II analysis of variance (ANOVA) was performed on the mixed-effects model. To test for MSI non-linearities, we calculated Miller’s Race Model Inequality (Miller, 1982) as described in Sinnett et al. (2008) as a benchmark.

#### 2.5.3. Alpha suppression

The frequency of interest (FOI) for the neuronal correlate of MSI was alpha (8-13 Hz). We expected to observe centro-parietal alpha suppression in the time window of interest (TOI), which was the first second after stimulus onset. To identify the region of interest (ROI) for the analysis, time-resolved oscillatory power was calculated for each subject each 0.02 s using a Short Time Fourier Transformation (frequencies of interest: 5 - 30 Hz, in steps of 1 Hz, in segments of 0.5s, Hanning window) for the interval of −1 to 2 s in Fieldtrip. Then, the time-frequency were averaged across subjects for each condition. The ROI was defined for all the subjects pooled together: CP1, CP3, P1, P3, P5.

We expected that alpha suppression in the AV condition would be larger than in the probabilistic sum of suppression in single modality A and V trials. Because linear operations, like AV – (A+V) commonly used for ERP contrasts, are not appropriate for power responses computed with fast Fourier or wavelet transformations, we followed the analysis pipeline suggested by Senkowski et al. (2007) and previously validated for complex stimuli by our group (Matyjek et al., 2024). First, we combined the amplitude of A-only and V-only trials linearly to create A+V trials. Then, we applied a Morlet wavelet transformation (with 7 cycles) to both multisensory (AV) and summed unisensory (A+V) epochs, converting them to the frequency domain. Then, the data were exported to R for further processing and analysis. Each epoch underwent absolute baseline correction relative to the mean value of the −300 to 0 ms period preceding the acoustic onset of the target word. Power values were then averaged across the ROI. Then, we performed bootstrapping on the A+V pairs, repeating the process 10,000 times to calculate their mean. Finally, to test prediction 3 (that AUT show a non-linear MSI effect in alpha suppression), we calculated z-scores of alpha power per participant (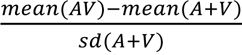), and tested whether they significantly deviated from 0 (with a one-sample t-test, as we expected a suppression effect). As pre-registered, we removed outliers defined as z-scores over or below the median +/-2x median absolute deviation.

To test whether AUT show smaller neural MSI effect than NT (prediction 4), we tested the difference between the z-scores in AUT and NT with a one-sided t-test.

#### 2.5.4. Exploratory analysis: spatio-temporal unfolding of MSI effects

Because literature on non-linear MSI effects in the frequency domain are scarce, an additional contribution of this article is an exploratory analysis of potential MSI effects that are complementary to the specific, pre-registered cluster and time window of interest of our planned alpha power analysis. For that, we extended the MSI analysis pipeline (described above for alpha) across spectral bands theta (4-7 Hz), alpha (8-13 Hz), beta (14-30 Hz), and gamma (30-48 Hz) across time windows. For theta and alpha we used windows of 0-0.5 s, and 0.5-1 s, and for beta and gamma, which are faster, we divided the available 1.5 s window into 3 bins of 300 ms (0-0.3 s, 0.3-0.6 s, 0.6-0.9 s). Each time window was absolute baseline-corrected using the mean amplitude value −0.3 to 0 pre-stimulus window. For wavelet transformations we used 5, 7, 8, and 10 cycles, for theta, alpha, beta, and gamma, respectively (see Matyjek et al., 2024 for validation of this approach using empirical and synthetic data).

## 3. Results

### 3.1. Behaviour: bimodal gain in accuracy

On average, AUT participants responded correctly in 96.5% of the AV trials, 86.8% of the V trials, and 72.1% of the A trials (see Figure 2, left panel). A logistic regression revealed a significant main effect of condition, *X^2^*(2)=78.79, *p*=.001. The odds of responding correctly in A trials were 94% smaller than in AV trials (1-0.06, *p*=.001, 95% CI [.04 .10]), and 78% smaller in V than in AV trials (1-0.22, *p*=.001, 95% CI [.14 .35]). Thus, prediction 1 was supported: AUT showed a behavioural effect of bimodal over unimodal speech. A model including the factor group (AUT, NT) showed a similar pattern of results: better accuracy in AV than A or V conditions, with no group differences or interaction of group and condition (see Figure 2, left panel, and Supplementary Material section 2).

**Figure 2.**
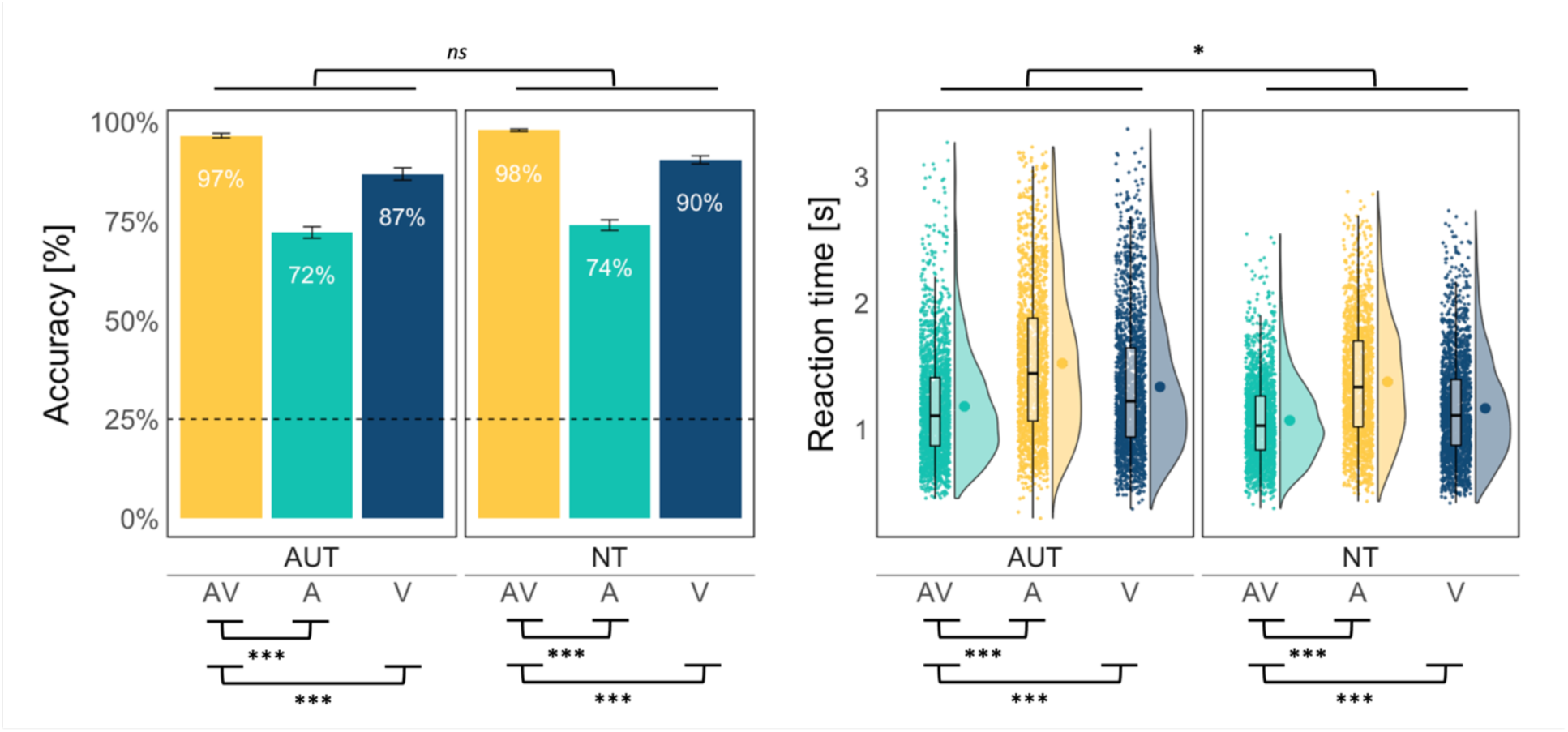
Mean accuracy (left panel) and reaction times (right panel) per group and condition. The error bars reflect standard error. The dashed line for accuracy marks the 25% chance performance in the task.

### 3.2. Behaviour: AUT vs. NT differences audio-visual gain

We compared the behavioural benefit (estimated as the difference in accuracy between AV and the largest of A or V) between AUT (mean=9.55) and NT (mean=7.37) with a t-test, which yielded no significant group effect, *t*(60.27)=1.38, *p*=.17 (see Figure 3, left panel). We additionally compared a model including the group factor against an intercept-only model, which yielded BF01 of 3.19. This suggests weak evidence in favour of the intercept-only model over the model including group.

**Figure 3.**
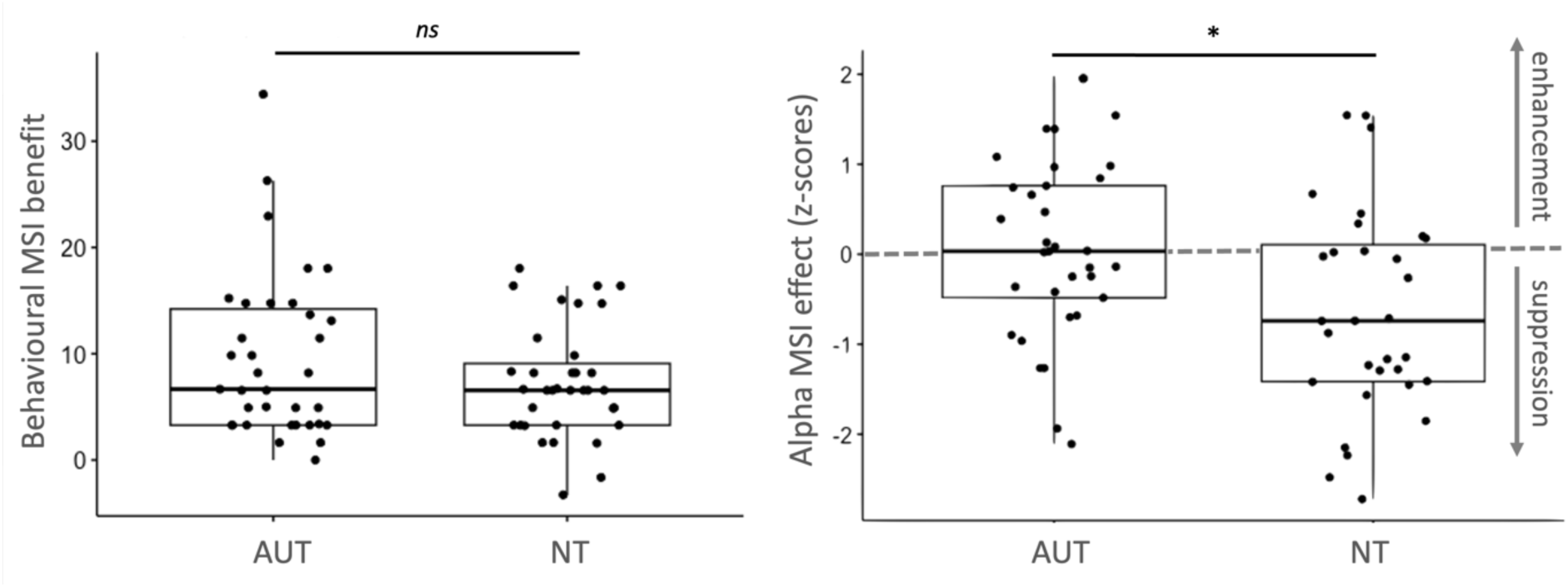
Behavioural benefit in word recognition (left panel) and neural MSI effect (z-scores) (right panel) across the groups.

### 3.3. Behaviour: exploratory analysis of reaction times

An ANOVA with the factors condition (AV, A, V) and group (AUT, NT) was performed using the GLMM yielded main effects of condition, *X^2^*(2)=1084.21, *p*<.001, and group *X*^2^(1)=5.51, *p*=.02. The interaction also reached significance, *X*^2^(2)=6.48, *p*=.04, but no pairwise comparisons of RTs between the groups survived correction for multiple comparisons and BF showed strong evidence in favour of a model without the interaction term, suggesting that this effect was negligible. Thus, RTs were faster for AV than V and for V than A, and this pattern held for both groups, although overall responses were faster in the NT than AUT group (see Figure 2, right panel). The distribution of observed RTs in AV trials never surpassed the theoretical race model with RTs in A and V trials in either group (see Supplementary Material section 3 for details).

### 3.4. Neural correlates: MSI effects on alpha suppression

In the alpha suppression analysis, we first checked for linear MSI effects (AV vs. A and AV vs. V) in AUT. A linear mixed model revealed a main effect of condition, *F*(2,6083.5)=9.43, *p*<.001, with stronger alpha suppression in AV compared to either A (*est*=117.93, *t*(6083.4)=4.31, *p*<.001) or V (*est=*71.61, *t*(6083.6)=2.62, *p*<.001). In a model including the factor group, condition was still significant, *F*(2,12271.1)=20.71, *p*<.001, but neither group nor the condition*group interaction were statistically significant (both *p*s>=.48; see Supplementary Material section 4). See TFRs per condition and group in Figure 4.

**Figure 4.**
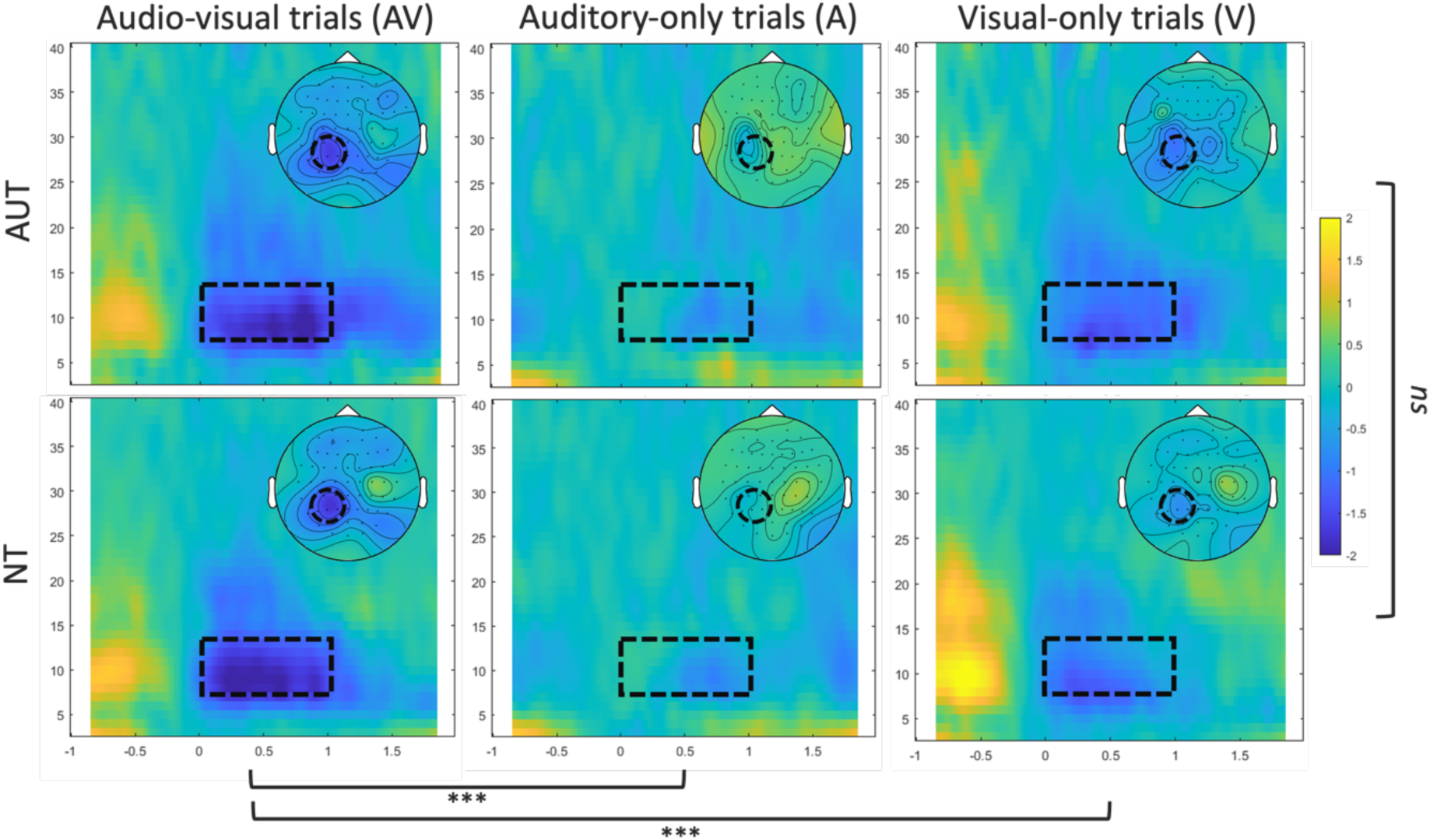
Time-frequency representations (TFR) per condition and group. The dashed rectangles mark the time (TOI; 0-1 s) and frequency of interest (FOI; alpha; 8-13 Hz). The TFRs are plotted for the region of interest in the alpha analysis (CP1, CP3, P1, P3, P5). The heads show the topography of the selected TOI and FOI and the dashed circles mark the electrodes in ROI.

To test for potential non-linear MSI effects in alpha suppression, we used z-scores of the deviation between observed mean alpha power in AV trials from the mean of the probabilistic sum of alpha power in A and V trials, with the contrast AV ≠ A+V. Contrary to prediction 3, we found no evidence for non-linear MSI effects in AUT, *t*(32)=0.29, *p*=.61. On the other hand, NT showed a significant non-linear effect, *t*(30)=-3.20, *p*=0.002.

### 3.5. Neural correlates: MSI effect difference between groups

To test whether AUT showed smaller alpha suppression MSI effect than NT (prediction 4), we tested the difference between the z-scores in AUT and NT with a one-sided t-test, which yielded significantly more negative z-scores (i.e., stronger suppression) in NT than in AUT, *t*(59.19)=2.65, *p*=.01 (see Figure 3, right panel).

### 3.6. Exploratory analysis: temporal and spatial unfolding of neural MSI effects

Figure 5 shows the MSI contrast extended to four frequency bands (theta, alpha, beta, gamma), across two (for theta and alpha) or three (for beta and gamma) time bins, in all electrodes. Although few points survived FDR correction (please note the number of comparisons performed: 61 electrodes x 2 or 3 bins = 122 or 183 comparisons per group and band) in this exploratory analysis (especially in the alpha band), these plots offer novel insights into the distribution of MSI effects in the brain in autistic and neurotypical adults and are therefore valuable for generation of future hypotheses.

### 3.7. Exploratory analyses: correlations between neural MSI, behavioural audio-visual gain, and individual traits

We additionally explored potential correlations between the behavioural benefit inaccuracy and alpha suppression across individuals (N=70). In short, there were no significant correlations either between the two outcomes, or between either of these and AQ, IQ, age, LSAS, or ADOS (in autistic participants with ADOS score; N=26), for either group or for all participants pooled together (see Supplementary Materials, section 5). Finally, we checked whether the groups significantly differed in their false positive and false negative rates for the catch trials. This was not the case (see Supplementary Materials, section 6).

## 4. Discussion

We investigated behavioural and neural correlates of multisensory integration of speech in autism, in a socially relevant complex environment. We tested autistic and neurotypical adult participants matched on age, sex, and non-verbal intelligence, all with average or above-average IQ and primarily communicating by speech. The key finding to emerge from this study is that while both groups benefited behaviourally to a similar extent from congruent multisensory information when performing a speech-in-noise comprehension task, the underlying neural processing mechanisms differed between the groups. As expected, both autistic and neurotypical groups showed greater alpha suppression during audio-visual speech perception than in either of the unisensory conditions. Yet, only the neurotypical group showed responses consistent with non-linear multisensory integration. Instead, the autistic group showed responses consistent with additive audio-visual interaction. This pattern of results suggests that while the autistic and neurotypical brains realise the MSI function differently, both result in efficient behavioural outcomes. This finding (1) extends previous results obtained in simpler, more constrained conditions to more ecologically valid conditions, where speech is embedded in situated linguistic, pragmatic, and social contexts; (2) furthers current MSI research on autistic populations to adulthood, a developmental stage that is both potentially informative and underrepresented in this line of research, and (3) considers behavioural and neural integration effects including group differences between linear and non-linear MSI processes. We discuss the implications of these findings.

### 4.1. Similar behavioural benefit in autistic and non-autistic groups

In line with our predictions (1 and 2), autistic participants showed improved speech-in-noise comprehension in the audio-visual (AV) compared to auditory-only (A) and visual-only (V) condition (similarly to neurotypicals). This was further supported by faster reaction times in AV than in A or V trials. Furthermore, the groups did not significantly differ from each other in the extent of this behavioural benefit. Previous studies showed that while autistic children and adolescents benefit less from multimodal speech information than their neurotypical peers (Brandwein et al., 2013; Foxe, Molholm, Del Bene, et al., 2015; Irwin et al., 2011; Smith & Bennetto, 2007; Stevenson et al., 2017), these differences wane with age (Beker et al., 2018; Foxe, Molholm, Del Bene, et al., 2015) and may vanish altogether in adults (Magnée et al., 2008). Thus, our data provide evidence to support the age-related catch-up process in multisensory gain by showing that adults in the autism spectrum generally benefit behaviourally from multisensory speech information to the same extent as neurotypicals.

At the same time, it is possible that the iconic gestures, more than other visual articulators, were particularly beneficial for speech-in-noise comprehension in this study. This is supported by the fact that the average accuracy rates for visual-only trials (89% correct) were larger than for auditory-only trials (73% correct) in both groups. Please note that most audio-visual speech studies report superior accuracy in the auditory modality compared to the visual, especially with large or unconstrained stimulus sets (e.g., Jaekl et al., 2015; Ross et al., 2007; Sumby & Pollack, 1954; for reviews, see Campbell, 2008; Navarra et al., 2012). Interestingly, these studies often exclude gestures or head movements. When this (admittedly artificial) constraint is lifted, this modality imbalance may revert to favour the visual channel, at least under certain circumstances. While there is substantial evidence for the role of gestures in communication (Kita & Emmorey, 2023; McNeill, 2000), little is known about how gestures and visual cues like lip movements interact with spoken language in naturalistic settings. This interplay is essential for understanding the general communication dynamics, and communication in autism in particular.

Although autism research has focused primarily on gesture production, leaving comprehension understudied (e.g., Dimitrova et al., 2017), autistic children understand gesture and speech similarly to their typically developing peers (Dimitrova et al., 2017; Dimitrova & Özçalışkan, 2022) and can benefit from watching iconic gestures in terms of focusing on the narrator and recalling the content (Dargue et al., 2021; Kurt, 2011). To date, only one other study has investigated the gestural benefit in a speech-in-noise task in autistic adults and, similarly to us, they found no difference in the size of the gestural enhancement between the groups (Mazzini et al., 2023). We used stimuli rich in visual articulators—such as lip movements, iconic gestures, and torso/face dynamics—to create a more naturalistic and integrated speech processing environment. As such, our study did not aim to isolate the specifically gestural enhancement from the overall multisensory benefit in speech processing. Regardless, the clear behavioural advantage of processing multimodal speech is evident in autistic adults and supports the previous (albeit scarce) literature.

### 4.2. Different neural MSI mechanisms between the groups

There are two main findings in our neural data. First, in terms of simple linear contrasts, we found significantly stronger alpha suppression in AV compared to A and V conditions in both autistic and non-autistic adults, with no group differences. Second, when looking at non-linear MSI interactions, calculated via the AV vs. A+V contrast, only the neurotypical group showed a significant effect. What is more, the size of the MSI effect was significantly larger in the neurotypical than the autistic group.

The AV vs. A+V contrast aims to determine whether the benefit from AV presentation could be accounted for by merely adding auditory and visual brain responses (Giard & Peronnet, 1999). Using this approach, two studies measured sensory event-related potentials (ERPs) to non-social, low-level stimuli found delayed and spatially limited MSI effects in autistic compared to non-autistic children and adolescents (Brandwein et al., 2013; Stefanou et al., 2020). In autistic children, this was accompanied by lesser behavioural benefit, but there were no behavioural group differences in adolescents (in line with the studies showing age-related improvements of behavioural MSI in autism; (Beker et al., 2018; Foxe, Molholm, Del Bene, et al., 2015)). In contrast, two other studies addressed MSI effects in ERPs for social stimuli in adult autistic and non-autistic participants (Magnée et al., 2008, 2011) and found no group differences in the AV vs. A+V (referred to as “lower-order MSI”). However, they also investigated “higher-order MSI”, defined as as congruency effects: short meaningless auditory utterances (/aba/ or /ada/) and (in)congruent visual speech videos (Magnée et al., 2008) or emotionally (in)congruent face-voice pairs (happy/fearful faces and laughing/gasping; (Magnée et al., 2011)). The congruency effect was observed in the neurotypical, but not autistic group, which the authors interpreted as an impairment in higher-order integration of complex information in autism.

Our results contrast the “lower-order MSI” effect in these studies. Arguably, our stimuli are more *complex*, including full words embedded in semantic, pragmatic, and social context, and multiple visual cues (in contrast to short non-semantic utterances and non-speech stimuli). Perhaps this complexity in our stimuli aligns better with the “higher-order” MSI effects reported by (Magnée et al., 2008, 2011). Regardless, our results extend previous findings to more naturalistic stimuli and contribute to the growing body of literature reporting neural MSI atypicalities in autism. Importantly, however, we emphasise that these atypicalities *cannot* be interpreted as an “impairment”. This is because of the clear lack of group differences between the groups in the behavioural benefit from multisensory information. When two brains achieve the same behavioural outcomes through different mechanisms, we can only describe these variations as differences rather than impairments.

### 4.3. Attention as a factor in MSI and its impact in autism

One explanation for the mismatch between behavioural and neural MSI correlates in our study is that autistic adults, unlike children, have developed compensatory strategies and brain mechanisms for integrating multisensory information, resulting in the absence of behavioural differences (at least those measured in this study) in adulthood. This is in line with the reported developmental delay in autistic children, estimated to be around six years ((Foxe, Molholm, Del Bene, et al., 2015); however, longitudinal studies are needed to confirm this). Yet, the specific nature of such compensatory mechanism remains to be elucidated.

For example, (Stefanou et al., 2020) suggested that higher-order attentional processes may activate MSI in autistic adolescents in later stages of processing, compensating for reduced early integration compared to neurotypicals. We cannot directly relate our data to these findings as ERPs used by Stefanou et al. (2020) operate on different temporal scales than oscillatory power dynamics used in this study (which are more appropriate for our more naturalistic stimuli; see a discussion in Matyjek et al., 2024). However, our exploratory analysis of the temporo-spatial unfolding of integration processes may generate future hypotheses in this regard. For instance, we observed that theta band activity might reveal significant MSI processes, with neurotypicals showing more integration effect than autistics. These differences were restricted to the second time window (0.5-1 second post-stimulus) (see Figure 5),which could support the proposition of later-stage processing playing a compensatory role in autism.

**Figure 5.**
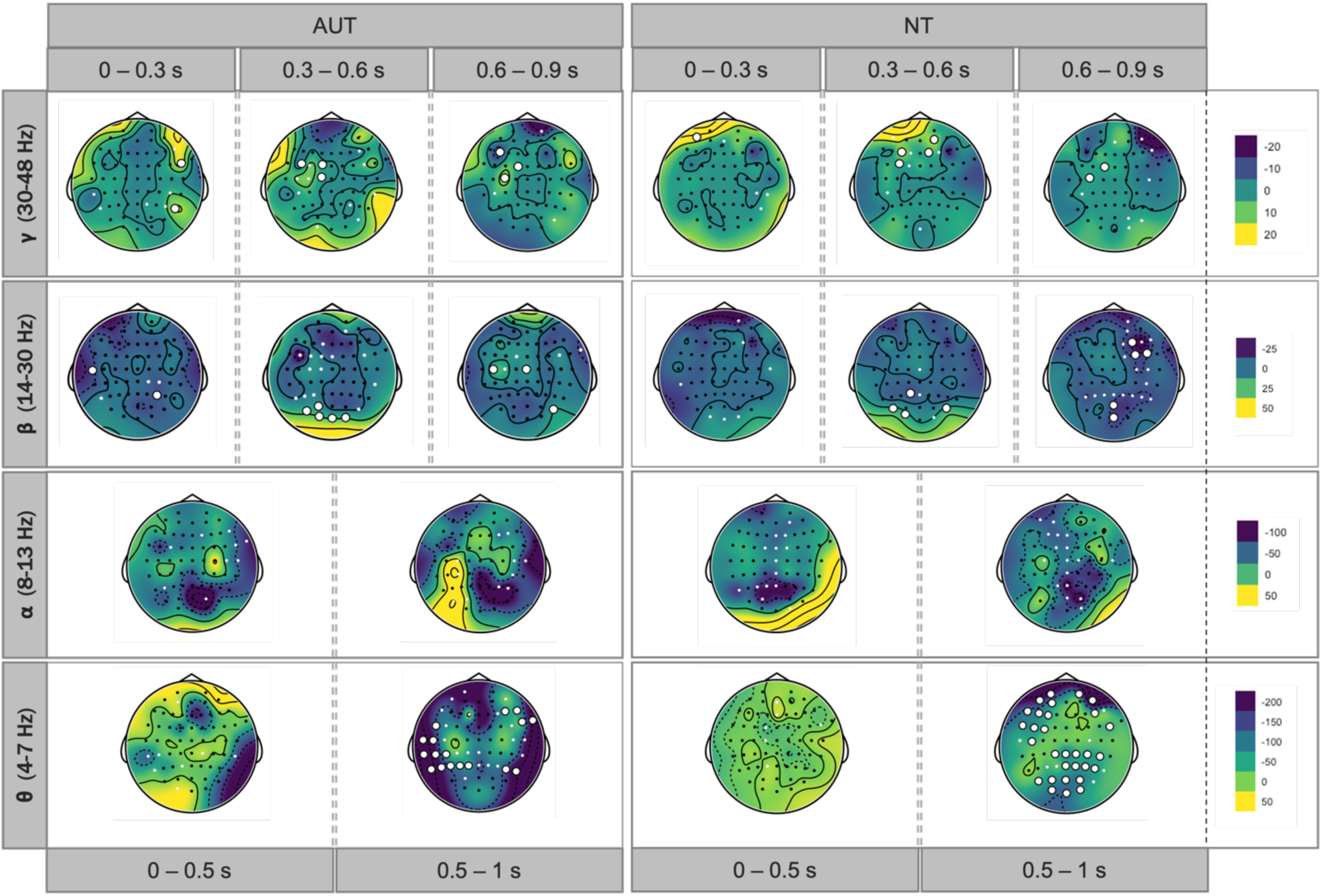
Spatio-temporal unfolding of MSI across bands. The colours of the topographical maps represent the difference of bimodal (AV) minus the sum of unimodal (A+V) mean power (*μ*V*²*). The circles correspond to the locations of the electrodes, with the large white circles marking the corrected MSI effect at this site and the small white circles representing additional sites with MSI effects which did not survive the FDR correction.

An alternative but related interpretation of our data concerns the differences in higher-order attentional processes between the groups. A good deal of evidence shows that MSI is subject to attentional demands, including the specific case of audio-visual speech integration (Alsius et al., 2005, 2007, 2014; Tiippana et al., 2004). Thus, brain correlated of audio-visual speech integration are mediated by sufficient attention to both modalities (Morís Fernández et al., 2015). However, autistic individuals seem to rely more on selective attention to auditory information instead of distributing attention across sensory channels, which could manifest in the same way as MSI impairments (Magnée et al., 2008; Marco et al., 2011).

Thus, MSI in autism may be preserved but dependent on attention. If attention engagement is not considered in the study design, observed differences in multisensory gain between groups may be wrongly interpreted as MSI deficits in autism. This is supported by electrophysiological studies suggesting that autistic individuals may need to actively engage attention in order to initiate early MSI, which is automatic in non-autistic subjects (Brandwein et al., 2013; Dunn et al., 2008; Magnée et al., 2008; Russo et al., 2010; Whitehouse & Bishop, 2008). For example, in a passive-viewing task, one study found an early (∼100 ms) MSI effect only in the neurotypical group and not in autism (Russo et al., 2010), while in an active task with explicit attentional demands, one study found equivalently early MSI effects in both groups (Brandwein et al., 2013). Similarly, (Magnée et al., 2011) manipulated attention and observed that autistic individuals showed typical brain responses in low-level MSI and in selective attention conditions, but not when divided attention was required by the task.. Thus, the lack of automatic early MSI in autism can be normalised through deployment of attention (Dunn et al., 2008) and/or can be compensated for by later attentional processes resulting in performance comparable to neurotypical controls (Stefanou et al., 2020).

In the present study, participants were required to divide their attention between audio and visual channels, as they were instructed to detect randomly appearing visual and auditory targets in ‘catch trials’. These targets were embedded in the naturalistic setting as ‘glitches,’ such as frozen frames and additional noise in the microphone. The autistic and neurotypical groups did not differ in their false alarm or miss rates to these catch trials. Although this ensures a minimal degree of attentional engagement across conditions in both groups, we cannot be certain that this manipulation effectively ensured the same degree of divided attention. The targets might have been sufficiently attention-grabbing to elicit reactions regardless of participants’ tendencies to use selective attention throughout the task. Future studies may need to manipulate attention more explicitly to calibrate precisely for divided attention.

Overall, it remains uncertain why autistic adults do not exhibit the same neural MSI as their neurotypical peers, despite clear behavioural benefits from multisensory speech information and similar differences in linear contrasts for neural responses between multisensory and unisensory trials. Differences in attention or strategy seem to be a promising avenue for further exploration. For example, autistic individuals may use attention-related compensatory neural mechanisms that results in behavioural benefits, or may rely on unisensory information instead of divided attention for audio-visual speech. These factors might underlie the additive integration processes which, at least in the context of this study, are sufficient to yield the behavioural advantage of audio-visual presentations.

### 4.4. No evidence of a link between MSI effects and social functioning in autism

Finally, we found no significant correlations between behavioural benefit and neural MSI, nor between these measures and quantified autistic traits or social functioning in the autism group (AQ and ADOS scores; see Supplementary Materials for details). It has been suggested that at least some of socio-communicative difficulties in autism may be *rooted* in atypical sensory processing and integration of multimodal information (Bahrick & Todd, 2012; Thye et al., 2018). There is a growing recognition and understanding of sensory processing in autism, with atypicalities in this domain proposed as potentially core characteristics and criteria for the diagnostic guidelines (American Psychiatric Association, 2013; Iarocci & McDonald, 2006; World Health Organization, 2021). In that vein, sensory atypicalities have been found to precede social cognition milestones in development (Estes et al., 2015; Robertson & Baron-Cohen, 2017), and to predict socio-communicative difficulties and eventual autism diagnosis (Turner-Brown et al., 2013). MSI differences have also been linked to difficulties in social and communicative skills in autism (Brandwein et al., 2015; Mongillo et al., 2008; Woynaroski et al., 2013). However, this was not observed in our study (please note that this null effect must be interpreted with caution, as our study was not designed to measure this potential relationship).

### 4.5. Limitations

First, we used pink noise to degrade speech comprehension, instead of socially-related noise (like various overlapping conversations). This may be relevant, given the frequently noted social processing difficulties in autism. For example, in a speech-in-noise task with various types of background speech-like noise, (Alcántara et al., 2004) observed that autistic adults performed worse than neurotypicals. However, in a population-based approach, (Tsuji & Imaizumi, 2024) did not find significant effects of social versus non-social noise despite generally diminished speech recognition in higher autistic traits. Examining this in clinically diagnosed samples remains important. Second, our conclusion about lack of differences in behaviour between autistic and neurotypical adults is based on the current statistical power of our design, and the measurement protocol used. These are comparable to previous studies; therefore, we believe we can confirm that behavioural MSI tends to equalise between these groups. Yet, some behavioural differences might appear, perhaps as a function of the different neural mechanisms detected, under other conditions with perhaps more challenging materials, or attentional requirements.

## 5. Conclusions

Our study investigated the behavioural and neural correlates of multisensory integration (MSI) in naturalistic speech among autistic and neurotypical adults. While both groups showed similar behavioural benefits from audio-visual speech, their neural processing mechanisms differed. Neurotypicals exhibited an interaction of auditory and visual information, whereas autistics showed additive processing of these inputs. Despite these neural differences, both groups achieved comparable behavioural outcomes, suggesting that the variations in neural mechanisms do not imply impairments. Attention may play a critical role, with autistics possibly relying on compensatory strategies developed over time. These findings extend our understanding of sensory processing and integration in autism, highlighting the importance of considering both behavioural and neural levels.

## Data Availability

The data and analysis code are available at https://osf.io/2b6s3/ (subproject: MSI in autistic vs. neurotypical adults).

## Acknowledgements

The authors extend their gratitude to the participants who volunteered for the study, sharing their valuable insights and comments. We also acknowledge and give our thanks to the clinical autism centres in Catalonia for their support in participant recruitment - special thanks are due to Autisme La Garriga.

## Ethics approval statement

The study protocol and data handling were approved by the Institutional Committee for Ethical Review of Projects at University Pompeu Fabra (CIREP-UPF; number 258). All participants gave a written consent to participate in the study.

## Data availability statement

The data and analysis code are available at https://osf.io/2b6s3/ (subproject “MSI in autistic vs. neurotypical adults”).

## Other permissions

All individuals depicted in the figures have provided written consent for the publication of their images in this scientific publication.

## Footnotes

1 The term ‘neurotypical’ refers to individuals with typical neurological development and functioning, but it often refers to individuals who are simply not autistic. This overlooks diversity within non-autistic populations. Nevertheless, in autism research, comparison groups are usually controlled for neurodivergent traits (e.g., anxiety) or clinical conditions (e.g., schizophrenia). Therefore, we use ‘neurotypical’ interchangeably with ‘non-autistic’ in this context.

2 We use identity-first language (“autistic person”) in this paper, as it is preferred by the majority of English-speaking individuals on the autism spectrum (e.g., Kenny et al., 2016), reduces stigma (Gernsbacher, 2017), and aligns with the preferences of all members of the autistic community directly or indirectly involved in this work.

