## Supplementary Material for "Multisensory Integration of Naturalistic Speech and Gestures in Autistic Adults"

### 1. Additional diagnostic information for AUT

| Participant | Diagnosis | Year of diagnosis | ADHD | Depression | Anxiety | Other diag. | Medication | Medication reason | ADOS-2 used | ADOS-2 total score | ADOS-2 comm. score | ADOS-2 social score | ADI-R used |
|---|---|---|---|---|---|---|---|---|---|---|---|---|---|
| 1 | ASD, I | 2019 | No | No | Yes | ADD | Decentan | mood regulator | Yes | 30 | - | - | No |
| 2 | ASD, I | 2021 | No | Yes | Yes | - | Paroxetina, Trazadana | depression | Yes | 9 | 4 | 4 | No |
| 3 | ASD, I | 2022 | No | No | Yes | - | Gabapetina | anxiety | Yes | 8 | 2 | 6 | Yes |
| 4 | ASD, I | 2016 | No | Yes | Yes | - | Escifalopram, Lorazepan | depression, anxiety | No | - | - | - | Yes |
| 5 | ASD, I | 2022 | No | Yes | Yes | - | - | - | Yes | 6 | 2 | 4 | No |
| 6 | ASD, I | 2013 | No | Yes | No | - | Citalopram | depression | No | - | - | - | No |
| 7 | ASD, I | 2022 | Yes | Yes | Yes | - | Concerta | ADHD | Yes | 7 | 2 | 5 | No |
| 8 | ASD, I | 2015 | No | Yes | No | - | Fluoxetina | depression | No | - | - | - | No |
| 9 | ASD, I | 2015 | Yes | No | Yes | PTSD | Brintellix, Lorazepam | depression, anxiety | Yes | 13 | 5 | 8 | No |
| 10 | ASD, I | 2021 | No | Yes | Yes | - | Diazepam | anxiety | Yes | 10 | 4 | 6 | Yes |
| 11 | ASD, I | 2023 | Yes | No | No | - | Concerta | ADHD | Yes | 13 | 5 | 8 | No |
| 12 | ASD, I | 2020 | No | No | No | ADD | Elvanse | ADD | Yes | 7 | 3 | 4 | No |
| 13 | ASD, I | 2021 | No | Yes | No | - | Escifalopram | depression | Yes | 10 | 6 | 4 | No |
| 14 | ASD, I | 2023 | No | Yes | Yes | - | - | - | Yes | 8 | 2 | 6 | No |
| 15 | ASD, I | 2022 | No | Yes | Yes | migraine | Fluoxetina | depression | Yes | 11 | 5 | 6 | No |
| 16 | ASD, I | 2017 | Yes | No | No | OCD | - | - | Yes | 7 | 2 | 5 | Yes |
| 17 | ASD, I | 2020 | No | Yes | Yes | - | Lorazepam, Paroxetina | depression, anxiety | Yes | 9 | 2 | 7 | Yes |
| 18 | ASD, I | 2022 | Yes | No | No | - | Elvanse | ADHD | Yes | 2 | 2 | 0 | No |
| 19 | ASD, I | 2023 | No | Yes | No | - | Sertralina | depression | Yes | 16 | 6 | 10 | No |

| | | | | | | | | | | | | | |
|---|---|---|---|---|---|---|---|---|---|---|---|---|---|
| 20 | ASD, I | 2019 | No | No | No | - | Carbamazepina | convulsions | Yes | 12 | 3 | 9 | Yes |
| 21 | ASD, I | 2022 | No | No | No | - | - | - | Yes | 7 | 2 | 5 | Yes |
| 22 | ASD, I | 2017 | No | Yes | Yes | - | - | - | Yes | 8 | 2 | 6 | No |
| 23 | ASD, I | 2021 | No | No | No | - | - | - | Yes | 9 | 4 | 5 | Yes |
| 24 | ASD, I | 2018 | No | No | No | - | - | - | Yes | 7 | 3 | 4 | No |
| 25 | ASD, I | 2022 | No | No | No | - | - | - | Yes | 15 | 5 | 10 | Yes |
| 26 | ASD, I | 2022 | No | Yes | Yes | - | Heipram | anxiety | No | - | - | - | Yes |
| 27 | ASD, I & II | 2023 | Yes | No | Yes | PTSD | - | - | Yes | 12 | 2 | 10 | Yes |
| 28 | ASD, I | 2023 | No | Yes | Yes | - | Fluoxetina | anxiety | Yes | 11 | 4 | 7 | Yes |
| 29 | ASD, I | 2023 | No | No | Yes | OCD | Citalopram | anxiety | No | - | - | - | No |
| 30 | ASD, I | 2022 | Yes | Yes | Yes | - | Concerta, Ribotril | ADHD, anxiety | No | - | - | - | Yes |
| 31 | ASD, I | 2020 | No | No | No | panic attacks | Risperdal, Noiafren | panic | No | - | - | - | No |
| 32 | ASD, I | 2023 | No | No | No | ADD, OCD | Sertralina | depression | No | - | - | - | No |
| 33 | Asperger's | 2008 | 1 | 0 | 0 | epilepsy | Vimpat (epilepsy), Strattera (ADHD), Sertralina (depression), Depakine (epilepsy), Quetiapina (schizophrenia), Selincro (alcohol) | epilepsy, ADHD, depression, schizophrenia, alcohol | No | - | - | - | No |
| 34 | TEA, I | 2022 | 0 | 0 | 1 | - | - | - | Yes | 10 | 3 | 7 | Yes |
| 35 | TEA, I | 2020 | 0 | 0 | 0 | - | - | - | Yes | 10 | 3 | 7 | No |

### 2. Accuracy in both groups

Average accuracy rates for groups and conditions are shown in Supp. Table 1. First, a logistic regression model with condition (AV, A, V) and group (AUT, NT) was built as follows:

```
glmer(formula = correct ~ cond * group + (1 + cond | ID) + (1 +
    cond | verb), data = DF, family = binomial(link = "logit"),
    control = glmerControl(optimizer = "bobyqa", optCtrl = list(maxfun = 200000)))
```

To estimate the global effects of its terms, we compared this model against models with the corresponding terms dropped using a likelihood test. This procedure revealed a statistically significant main effect of condition, $X^2(2) = 127.61$, $p < .001$, so that accuracy in AV was larger than in A and larger than in V. The odds of responding correctly in the A and V conditions were, respectively, 94% (1-0.06, 95% CI [.04 .10]) and 75% (1-0.25, 95% CI [.17 .40]) significantly (both *p*s < .001) smaller than in the AV condition. Interaction and group effects were not statistically significant (respectively, $X^2(2) = 3.40$, $p = .18$ and $X^2(2) = 1.61$, $p = .20$).

Thus, both groups showed the behavioural MSI effect in the accuracy rates of word detection, so that participants were more accurate in bimodal than in unimodal trials. The groups did not significantly differ in their responses in general or for different conditions.

**Supp. Table 1: Average accuracy rates in word detection by group and condition.** SDs are reported in brackets.

| Group | AV | A | V |
|---|---|---|---|
| **AUT** | 96.53% (3.55) | 72.11% (8.56) | 86.84% (9.13) |
| **NT** | 97.88% (1.77) | 73.95% (7.84) | 90.42% (5.9) |
| **All** | 97.21% (2.87) | 73.03% (8.2) | 88.63% (7.84) |

### 3. Reaction times in both groups

Mean RTs per group and condition are shown in Supp. Table 2.

**Supp. Table 2: Average reaction times by group and condition.** SDs are reported in brackets.

| Group | AV | A | V |
|---|---|---|---|
| **AUT** | 1.18s (0.43) | 1.52s (0.59) | 1.34s (0.53) |
| **NT** | 1.07s (0.32) | 1.38s (0.46) | 1.16s (0.4) |
| **All** | 1.12s (0.38) | 1.45s (0.53) | 1.25s (0.48) |

To analyse RTs, which do not follow normal distribution, we first built two models with Gamma distribution and Inverse Gaussian distribution, and the same fixed and random structure:

```
(1) glmer(RT ~ cond*group + (1|ID) + (1|verb), data = RT,
    family=Gamma(link="identity"))

(2) glmer(RT ~ cond*group + (1|ID) + (1|verb), data = RT,
    family=inverse.gaussian(link="identity"))
```

Then, we compared models 1 and 2 for AIC and BIC, which were both lower for model 1 ($AIC_1$ = 9398.974, $AIC_2$ = 9556.402, $BIC_1$ = 9465.208, $BIC_2$ = 9622.636). This model yielded main effects of condition, $X^2(2)$=1084.21, $p$<.001 and group $X^2(1)$=5.51, $p$=.02, and their interaction, $X^2(2)$=6.48, $p$=.04. However, no pairwise comparisons of RTs between the groups survived correction for multiple comparisons (Supp. Table 3). Additionally, the BF01 was 454.43, suggesting strong evidence in favour of a model without the interaction effect..

Together, RTs were faster for AV than V and for V than A, and NT showed faster RTs than AUT, but the groups responded similarly across conditions (**Supp. Fig. 1**). The distribution of observed RTs in AV trials never surpassed the race model with RTs in A and V trials in either group (Supp. Fig. 2).

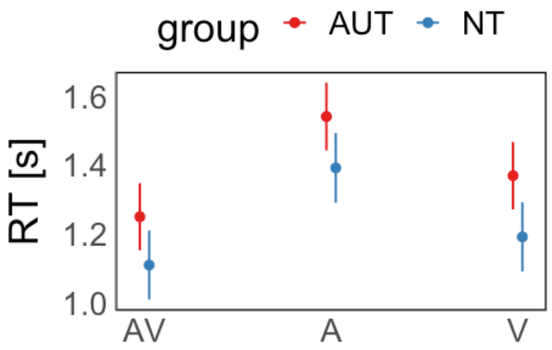

**Supp. Fig. 1: Predicted reaction times for groups and conditions.** Error bars mark 95% CI.

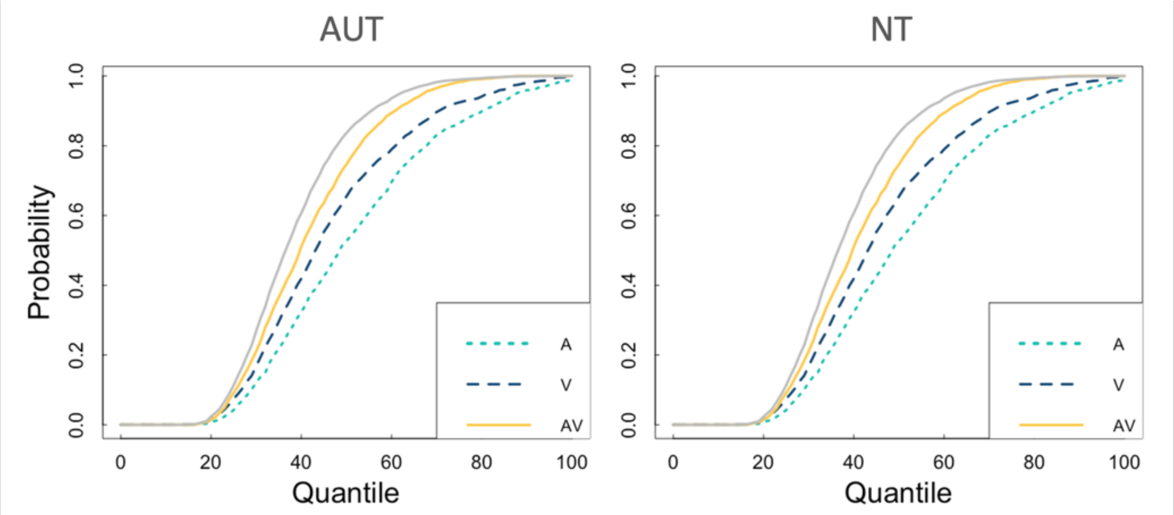

**Supp. Fig. 2: Representation of the race model test for each group.**

**Supp. Table 3: Pairwise comparisons for the effect of condition and group interaction on RTs (with Holm correction).**

| Contrast | Est. | Std. Error | z value | $p_{corr}$ |
|---|---|---|---|---|
| **AUT - NT** | | | | |
| $NT_A$ - $AUT_A$ | -0.149 | 0.067 | -2.216 | 0.163 |
| $NT_{AV}$ - $AUT_{AV}$ | -0.140 | 0.067 | -2.107 | 0.206 |
| $NT_V$ - $AUT_V$ | -0.178 | 0.067 | -2.664 | 0.052 |
| **AUT cond** | | | | |
| $AUT_A$ - $AUT_V$ | 0.172 | 0.013 | 12.852 | 0.000 |
| $AUT_{AV}$ - $AUT_A$ | -0.291 | 0.013 | -22.851 | 0.000 |
| $AUT_{AV}$ - $AUT_V$ | -0.119 | 0.011 | -10.725 | 0.000 |
| **NT cond** | | | | |
| $NT_A$ - $NT_V$ | 0.201 | 0.012 | 16.285 | 0.000 |
| $NT_{AV}$ - $NT_A$ | -0.283 | 0.012 | -23.738 | 0.000 |
| $NT_{AV}$ - $NT_V$ | -0.082 | 0.010 | -8.060 | 0.000 |

### 4. Linear and non-linear MSI effects in alpha suppression for both groups

#### Linear MSI in both groups

We tested linear MSI (AV vs. A and AV vs. V) in both groups by building a linear regression mixed model with condition (AV, A, V), group (AUT, NT), and their interaction as predictors of alpha suppression (with random intercepts for participants). This analysis yielded a main effect of condition, $F(2,12271.1)=20.71$, $p<.001$, with alpha more suppressed in AV than A (*est.*=106.60, $p_{corr}<.001$) and in AV than V (*est.*=79.38, $p_{corr}<.001$). There were no statistically significant effects of group, $F(1,70.3)=0.51$, $p=.48$, BF01 = 86.01, or condition*group interaction, $F(2,12271.1)=0.62$, $p=.54$, BF01 = 666.72.

#### Non-linear MSI in both groups

Contrary to our prediction 3, we found no evidence for non-linear MSI effects in AUT, tested with a one-sided t-test of z-scores against 0, $t(32) = 0.29$, $p=.61$. As pre-registered, these z-scores were free of outliers (i.e., values over or below the median +/-2x median absolute deviation). The same test on all the z-scores (including outliers) was also insignificant, $t(34) = -0.45$, $p=.33$. The same test for NT showed a significant result on z-scores without the outliers, $t(30) = -3.20$, $p = 0.002$ (but not when including the outliers, t(34) = -1.31, $p = 0.1$).

### 5. Secondary analysis: correlations

**We additionally explored possible correlations between our main behavioural and neural outcomes: the behavioural benefit from multisensory information (accuracy rate for AV – max(A,V)), and the z-score for the AV – (A+V) contrast in alpha suppression. All correlations are shown in**

**Supp. Fig. 3. In short, there were no significant correlations either between the two MSI outcomes, or between these and AQ, IQ, age, LSAS, or ADOS (taking only the autistic participants who had an ADOS score; N=26), for either group or for all participants together (see**

**Supp. Fig. 3 panel A). After removing the z-score outliers, the neural MSI (z-scores) correlated with autistic and social anxiety traits, but only across all subjects (not within groups; see**

Supp. Fig. 3 panel B). We stress that these are exploratory correlations on a small number of data points, with no corrections for multiple comparisons. Thus, we consider them not significant in the light of this study.

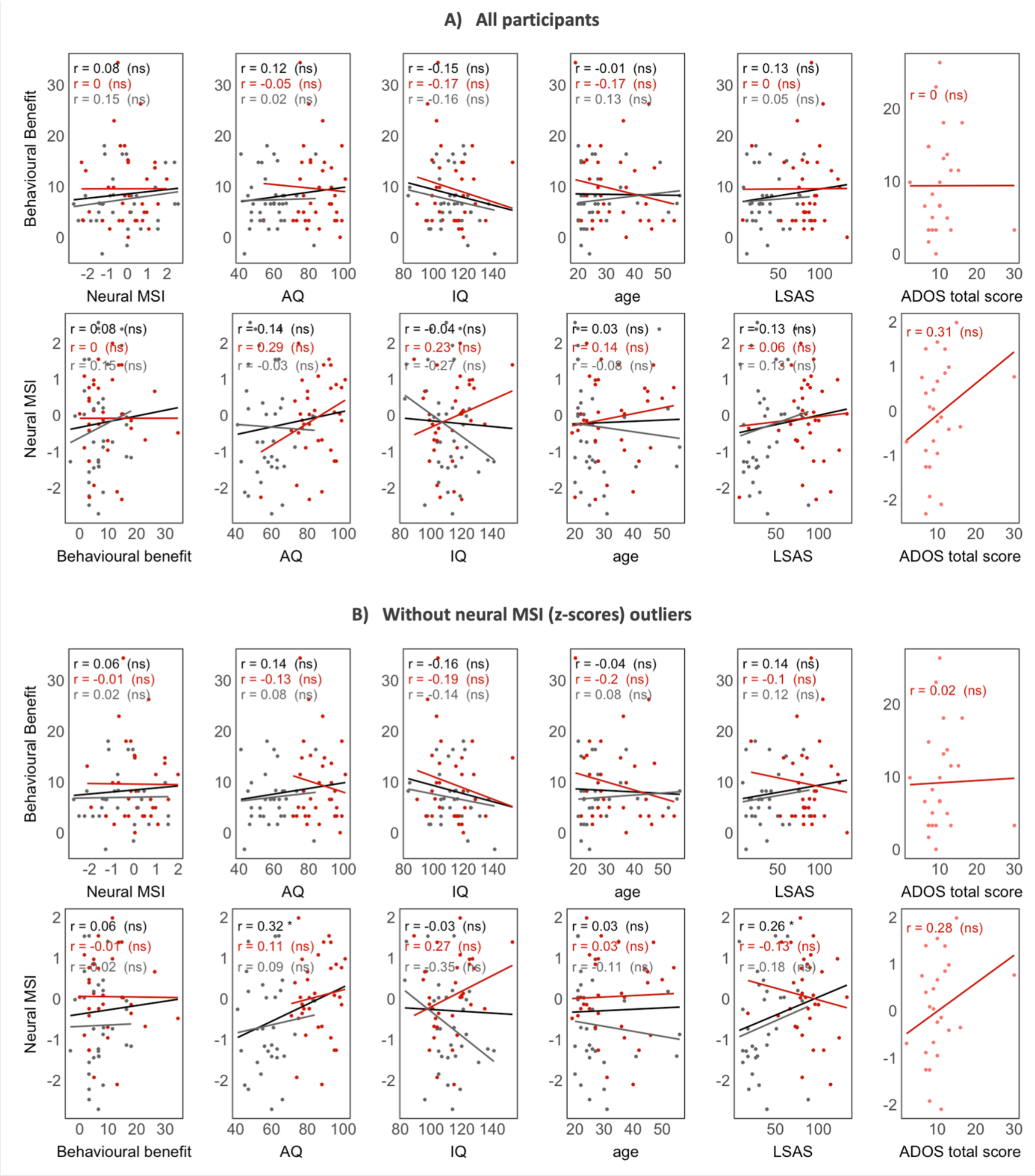

**Supp. Fig. 3 Exploratory correlations between the behavioural and neural MSI outcomes and AQ, IQ, age, LSAS, and ADOS (for AUT participants only).**

### 6. Secondary analysis: responses to targets in catch trials

To ensure that participants paid attention to the task in both sensory channels, participants were asked to monitor for occasional glitches (catch trials) in either the visual or the auditory channel (frozen frames or a burst of noise in the microphone, respectively). There were 9 catch trials in the auditory and 10 in visual modality. The mean false positive and false negative responses per group are shown in Supp. Table 4.

**Supp. Table 4 Mean false positive and false negative responses per group.** FP = false positive, FN = false negative. For FP, AV/A/V signify the condition of the trial. For FN, A/V signify the modality in which the target was presented. SDs are reported in brackets.

| Group | FP in AV | FP in A | FP in V | FN in A | FP in V | FP | FN |
|---|---|---|---|---|---|---|---|
| **AUT** | 0.43 (0.9) | 2.57 (10.10) | 3.06 (6.61) | 0.89 (1.4) | 0.89 (1.105) | 6.06 (16.72) | 1.77 (1.78) |
| **NT** | 0.14 (0.5) | 1.89 (6.11) | 0.31 (0.80) | 0.94 (1.37) | 0.40 (0.65) | 2.34 (6.11) | 1.34 (1.51) |
| **All** | 0.29 (0.73) | 2.23 (8.30) | 1.69 (4.87) | 0.91 (1.39) | 0.64 (0.93) | 4.2 (12.64) | 1.56 (1.66) |

Some autistic participants (N=5) and one neurotypical participant exhibited particularly high (>10 across conditions) false positive rates (i.e., reporting a target when there was none; see the code/html file with all results), which was due to not understandings of the instructions specific to the targets (even after passing the training). After receiving additional explanations from the experimenter during the breaks between blocks, these participants significantly reduced their false positives. Regardless of including or excluding these participants, there were no significant differences in false positive, of false negative rates between the groups (for all participants, FP: $t(42.93) = 1.23$, $p = .22$, FN: $t(66.25) = 1.08$, $p = .28$).
